# Association between frequency of rehabilitation therapy and long-term mortality after stroke : a nationwide cohort study

**DOI:** 10.1101/2023.12.05.23299564

**Authors:** Je Shik Nam, Seok-Jae Heo, Yong Wook Kim, Sang Chul Lee, Seung Nam Yang, Seo Yeon Yoon

## Abstract

**Background:** Poststroke rehabilitation reportedly improves functional outcomes and minimizes disability. However, previous studies have demonstrated conflicting results regarding the effects of rehabilitation therapy on post-stroke mortality. Therefore, we aimed to investigate the association between rehabilitation therapy within the first six months after stroke and long-term all-cause mortality in patients with stroke using data from the Korean National Health Insurance System.

**Methods:** A total of 10,974 patients newly diagnosed with stroke using ICD-10 codes (I60-I64) between 2013 and 2019 were enrolled and followed-up for all-cause mortality until 2019. Post-stroke patients were categorized into three groups according to the frequency of rehabilitation therapy: no rehabilitation therapy, ≤ 40 sessions, and > 40 sessions. Cox proportional hazard models were used to assess the mortality risk according to rehabilitation therapy stratified by disability severity.

**Results:** Higher frequency of rehabilitation therapy was associated with significantly lower post-stroke mortality in comparison to no rehabilitation therapy (HR=0.88, 95% CI 0.79-0.9 9), especially among individuals with severe disability after stroke (HR=0.74, 95% CI 0.62-0.87). An inverse association between number of rehabilitation therapy sessions and mortality was identified in a multivariate Cox regression model with restricted cubic splines. In the context of stroke type, higher frequency of rehabilitation therapy was associated with reduced mortality rates compared to no rehabilitation therapy only in patients with hemorrhagic stroke (HR=0.60, 95% CI 0.49-0.74). While socioeconomic factors were not associated with mortality, older age, male sex, and pneumonia were associated with increased mortality risk, regardless of disability severity.

**Conclusions:** Post-stroke rehabilitation therapy within six months of stroke onset seems to play a substantial role in reducing long-term mortality after stroke. A higher frequency of rehabilitation therapy is recommended for post-stroke patients, particularly among those with severe disability.

## Introduction

Stroke is the second-leading cause of death and a major cause of disability worldwide.^1^ In Korea, cerebrovascular diseases were the fourth leading cause of death, accounting for 7.1% of all deaths in 2021.^2^ The increase in incidence of stroke by 29.7% coupled with decrease in mortality rates by 12.8% from 2014 to 2019 highlights the importance of comprehensive approach in long-term stroke management, including rehabilitation treatment.^3^

Most stroke survivors suffer from long-term sequelae, such as motor impairment, cognitive deficits^4^, and neuropsychiatric symptoms, including depression and anxiety.^5^ These factors affect patient’s mobility, activities of daily living, and societal participations, resulting in low quality of life and burden on the healthcare system.^4, 6-8^Several studies have demonstrated that effective rehabilitation minimizes these disabilities and helps functional recovery of post-stroke survivors, which not only increases their satisfaction but also decreases the socioeconomic costs.^9, 10^ It can also lead to substantial improvement in activities of daily living and functional level with increased intensity.^11, 12^ Moreover, reduction in disability and improvement of physical and social functions through rehabilitation has even been reported in patients with chronic stroke. ^13^

It is well known that acute stroke management in stroke care unit significantly reduces stroke-associated mortality.^14, 15^ Some studies have shown positive effects of rehabilitation therapy on mortality in patients with stroke.^16, 17^ On the other hand, a non-significant association between these two factors has also been reported.^18, 19^ Previous studies have examined the impact of rehabilitation treatment received within the initial three months, ^16, 20^ one year, ^21^ and two years ^18^ on long-term mortality. In this study, we evaluated the effect of rehabilitation therapy received within six months after stroke onset on mortality, as the recent rehabilitation treatment paradigm has shifted towards maximizing functional recovery after stroke, emphasizing early treatment and maintaining adequate treatment for the first six months after stroke.^22, 23^ Therefore, this study aimed to investigate the association between rehabilitation therapy received within six months of stroke onset and long-term all-cause mortality in post-stroke patients using large nationwide cohort data. We also evaluated the associations according to severity of disability, stroke type, and sex to evaluate whether the association between rehabilitation treatment and mortality remained consistent

## Methods

### Data source

The Korean National Health Insurance Service (NHIS) is a mandatory health insurance coverage provided by the government to all Korean citizens. Each medical institution electronically submits all healthcare utilization data to the NHIS for reimbursement purposes; therefore, information on healthcare utilization in Korea is centralized in the NHIS database. The Korean NHIS – National Sample Cohort data is a representative sample database containing information on approximately one million Koreans. The data includes a unique anonymous number for each patient; summary of sociodemographic factors, such as age, sex, type of insurance; and medical information, such as a list of diagnoses according to the International Classification of Diseases (ICD-10) along with the procedural codes. This study was approved by the Institutional Review Board of the Korea University Guro Hospital, which waived the need for informed consent. The study was performed in accordance with the tenets of Declaration of Helsinki.

### Study population

We selected patients who were newly diagnosed with stroke, defined as primary diagnosis according to ICD-10 codes (I60-I64) between 2003 and 2019. We only included patients admitted to the hospital with a diagnosis of stroke based on brain imaging evaluations, such as magnetic resonance imaging or computed tomography. Subsequently, we excluded patients (1) with a combined diagnosis of transient ischemic attack, (2) who had already been registered in the National Disability Registration (NDR) system for brain impairment before the diagnosis of stroke, and (3) who died within six months of stroke onset. Finally, 10,974 patients with new-onset stroke were enrolled and followed-up for all-cause mortality until 2019.

### Rehabilitation therapy

In Korea, various rehabilitation treatments such as physical therapy, occupational therapy, dysphagia therapy, cognitive therapy, and speech therapy have been implemented to recover from impairment after stroke. This study focused on “rehabilitative developmental therapy for central nervous system disorders (claim code: MM105), which is the most important therapy for functional recovery after stroke in Korea. This therapy can be claimed when a qualified rehabilitation specialist or physical therapist performs 1:1 professional individualized patient-centered rehabilitation therapy for patients with central nervous system injuries for more than 30 min. It can be performed twice a day for the first two years after stroke onset, and then only once a day thereafter. In this study, median value of the number of cumulative rehabilitation therapy within six months after stroke onset was 40 (Interquartile range 9.0-99.5) among patients who received at least one rehabilitation therapy. Therefore, patients with stroke were classified into three groups on the basis of frequency of rehabilitation therapy: ‘no rehabilitation therapy’, ‘rehabilitation therapy ≤ 40 sessions’, and ‘rehabilitation therapy > 40 sessions’.

### Other variables

The endpoint of this study was death of patients with stroke. All-cause mortality up to December 31, 2019, was evaluated. Ages were categorized into four groups: < 60 years, 60-69 years, 70-79 years, and ≥80 years. Areas of residence were categorized into capital, urban, and rural. The National Health Insurance premium was used as a proxy measure of income as it is proportional to monthly income, including earnings and capital gains. NDR from brain impairment was extracted and used as a proxy for disease severity. The Korean government classifies NDR from brain impairment into six grades according to activities of daily living based on the Modified Barthel Index (MBI). We classified NDR grades into the following three categories: none (97 ≤ MBI), mild to moderate (NDR 4-6 grades, 70 ≤ MBI ≤ 96), and severe (NDR 1-3 grades, MBI < 70). Comorbidity was defined using the Charlson Comorbidity Index (CCI). In this study, cerebrovascular disease and hemiplegia were excluded when calculating the CCI score because these two diagnoses were the main characteristics of the participants in this study and not comorbidities.

### Statistical analysis

Baseline demographic and medical data were presented as numbers (percentages) according to rehabilitation therapy. The variables in the three groups were compared using the chi-square test. The primary outcome was all-cause mortality according to rehabilitation therapy in patients with stroke. Cox proportional hazard models were used to assess the mortality risk according to rehabilitation therapy using three models with adjustments for confounding variables. Survival plots were constructed and compared using log-rank tests. In addition, a restricted cubic spline function was used to investigate the dose-response association between the number of cumulative rehabilitation therapies and mortality. Subgroup analyses were performed according to disability severity, stroke type, and sex to evaluate whether the association between rehabilitation treatment and mortality remained consistent. All statistical analyses were performed using SAS, version 9.4 software (SAS Institute Inc), and a two-sided p < 0.05 was considered statistically significant.

## Results

### Characteristics of the study participants

Table 1 presents the characteristics of the 10,974 post-stroke patients included in this study. When categorizing the patients into three groups based on the frequency of rehabilitation sessions, there were 6,738 patients in the group without rehabilitation therapy, 2,122 in the group with ≤40 sessions, and 2,114 in the group with >40 sessions. During the follow-up duration (mean 4.85 ± 3.35 years), among the 10,974 patients with stroke, 3,103 (28. 3%) died.

**Table 1.** Characteristics of the study participants.

| Variables | Rehabilitation therapy |  |  | p-value |
| --- | --- | --- | --- | --- |
| | No | Yes ($\leq 40$) | Yes ($> 40$) | |
| **N** | 6738 | 2122 | 2114 |  |
| **Age (years)** |  |  |  | <.001 |
| < 60 | 1846 (27.3) | 548 (25.8) | 656 (31.0) |  |
| 60-69 | 1406 (20.9) | 460 (21.7) | 470 (22.2) |  |
| 70-79 | 1817 (27.0) | 556 (26.2) | 543 (25.7) |  |
| $\geq 80$ | 1669 (24.8) | 558 (26.3) | 445 (21.1) | |
| **Sex** |  |  |  | 0.294 |
| Male | 3641 (54.0) | 1123 (53.0) | 1104 (52.2) |  |
| Female | 3097 (46.0) | 999 (47.0) | 1010 (47.8) |  |
| **Income levels** |  |  |  | 0.031 |
| Q1 (lowest) | 1219 (18.1) | 347 (16.3) | 331 (15.7) |  |
| Q2 | 1267 (18.8) | 423 (20.0) | 457 (21.6) |  |
| Q3 | 1617 (24.0) | 509 (24.0) | 509 (24.1) |  |
| Q4 (highest) | 2635 (39.1) | 843 (39.7) | 817 (38.7) |  |
| **Area of residence** |  |  |  | <.001 |
| Capital | 1085 (16.1) | 328 (15.5) | 380 (18.0) |  |
| Urban | 1480 (22.0) | 558 (26.3) | 563 (26.6) |  |
| Rural | 4173 (61.9) | 1236 (58.2) | 1171 (55.4) |  |
| **Insurance type** |  |  |  | <.001 |
| National health insurance | 6176 (91.7) | 1994 (94.0) | 1968 (93.1) |  |
| Medical aid | 562 (8.3) | 128 (6.0) | 146 (6.9) |  |
| **Disability** |  |  |  | <.001 |
| None | 5088 (75.5) | 1389 (65.5) | 598 (28.3) |  |
| Mild to moderate | 914 (13.6) | 403 (19.0) | 530 (25.1) |  |
| Severe | 736 (10.9) | 330 (15.5) | 986 (46.6) |  |
| **Stroke subtype** |  |  |  | <.001 |
| Ischemic | 4573 (67.9) | 1443 (68.0) | 1206 (57.0) |  |
| Hemorrhagic | 1964 (29.1) | 672 (31.7) | 898 (42.5) |  |
| Not specified | 201 (3.0) | 7 (0.3) | 10 (0.5) |  |
| **Charlson comorbidity index** |  |  |  | 0.498 |
| 0 | 57 (0.9) | 14 (0.6) | 9 (0.4) |  |
| 1 | 128 (1.9) | 38 (1.8) | 39 (1.8) |  |
| 2 | 292 (4.3) | 95 (4.5) | 82 (3.9) |  |
| $\geq 3$ | 6261 (92.9) | 1975 (93.1) | 1984 (93.9) | |
| **Other co-morbidities<sup>a</sup>** |  |  |  |  |
| Hypertension | 6161 (91.4) | 2006 (94.5) | 2025 (95.8) | <.001 |
| Dyslipidemia | 6174 (91.6) | 1983 (93.5) | 1983 (93.8) | <.001 |
| Pneumonia | 3496 (51.9) | 1102 (51.9) | 1199 (56.7) | <.001 |
| Urinary tract infection | 2943 (43.7) | 984 (46.4) | 1305 (61.7) | <.001 |

In the group with higher frequency of rehabilitation therapy, the patients were younger (p<0.001) and showed lower proportion of lowest income level (p=0.031) and rural residence (p<0.001). This group had more number of severely disabled stroke patients than their counterparts (p<0.001). In the context of stroke type, there was a greater proportion of patients with hemorrhagic stroke in the higher frequency rehabilitation therapy group than in the other groups (p<0.001).

### Mortality rates according to frequency of rehabilitation therapy

Table 2 shows the hazard ratio (HR) for all-cause mortality according to the frequency of rehabilitation therapy based on Cox proportional hazard models. In patients with stroke, compared to no rehabilitation therapy group, the group with higher frequency of rehabilitation therapy revealed significantly lower mortality rates (HR=0.88, 95% confidence interval [CI] 0.79-0.99). We also investigated the association between rehabilitation therapy and post-stroke mortality according to disease severity. The results showed that the mortality rate was lower in non-disabled patients who received <40 rehabilitation therapy sessions after stroke (HR = 0.88, 95% CI 0.77-0.99). Mortality rates were not associated with the frequency of rehabilitation therapy in stroke patients with mild to moderate disabilities. For severely disabled patients after stroke, patients who received >40 rehabilitation therapy sessions showed significantly lower mortality rates than those without rehabilitation therapy (HR=0.74, 95% CI 0.62-0.87). The results of the multivariable-adjusted survival curves and log-rank tests for mortality according to the frequency of rehabilitation therapy in patients with stroke are displayed in Figure 1.

**Figure 1.**
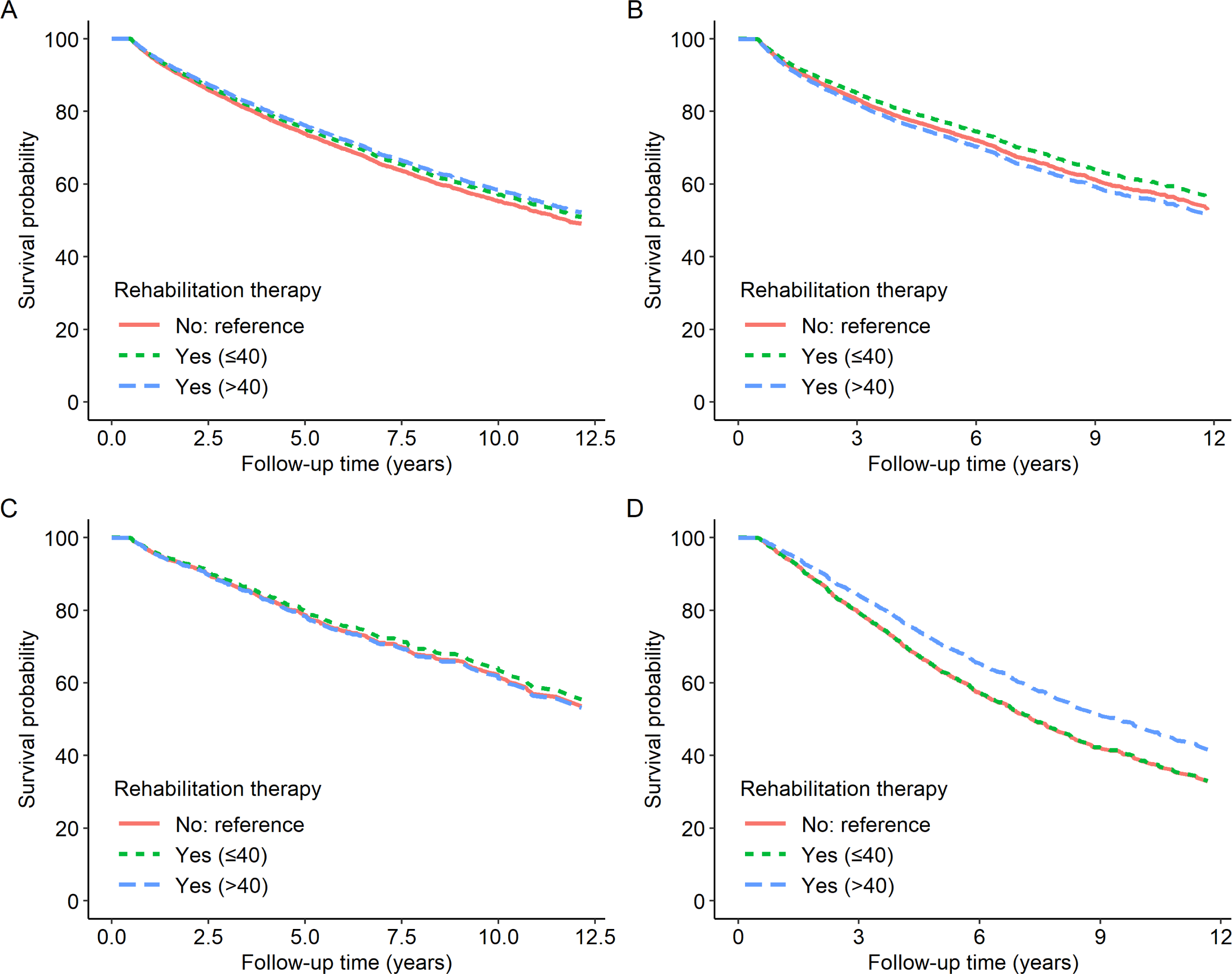
Multivariable-adjusted survival curves and log-rank tests for all-cause mortalit y according to frequency of rehabilitation therapy in patients with stroke. (A) Total, (b) No disability, (c) mild to moderate disability, and (d) severe disability.

**Table 2.**
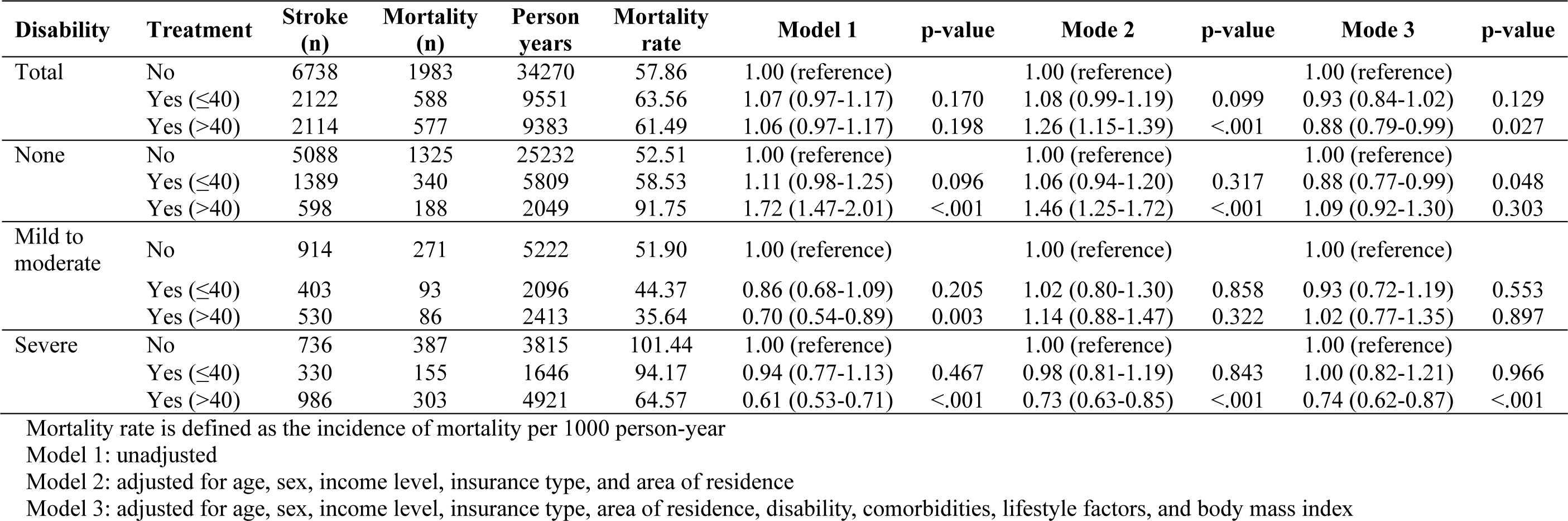
All-cause mortality by rehabilitation therapy frequency in patients with stroke stratified by disability severity moderate.

In multivariable Cox regression models with restricted cubic splines, the number of rehabilitation therapy sessions revealed an inverse association with mortality rate, showing a gradual decrease in the mortality rate in patients who received more than 60 sessions of rehabilitation therapy (Figure 2). In particular, in stroke patients with severe disability, an inverse dose-response association was found between number of rehabilitation therapy sessions and mortality rate.

**Figure 2.**
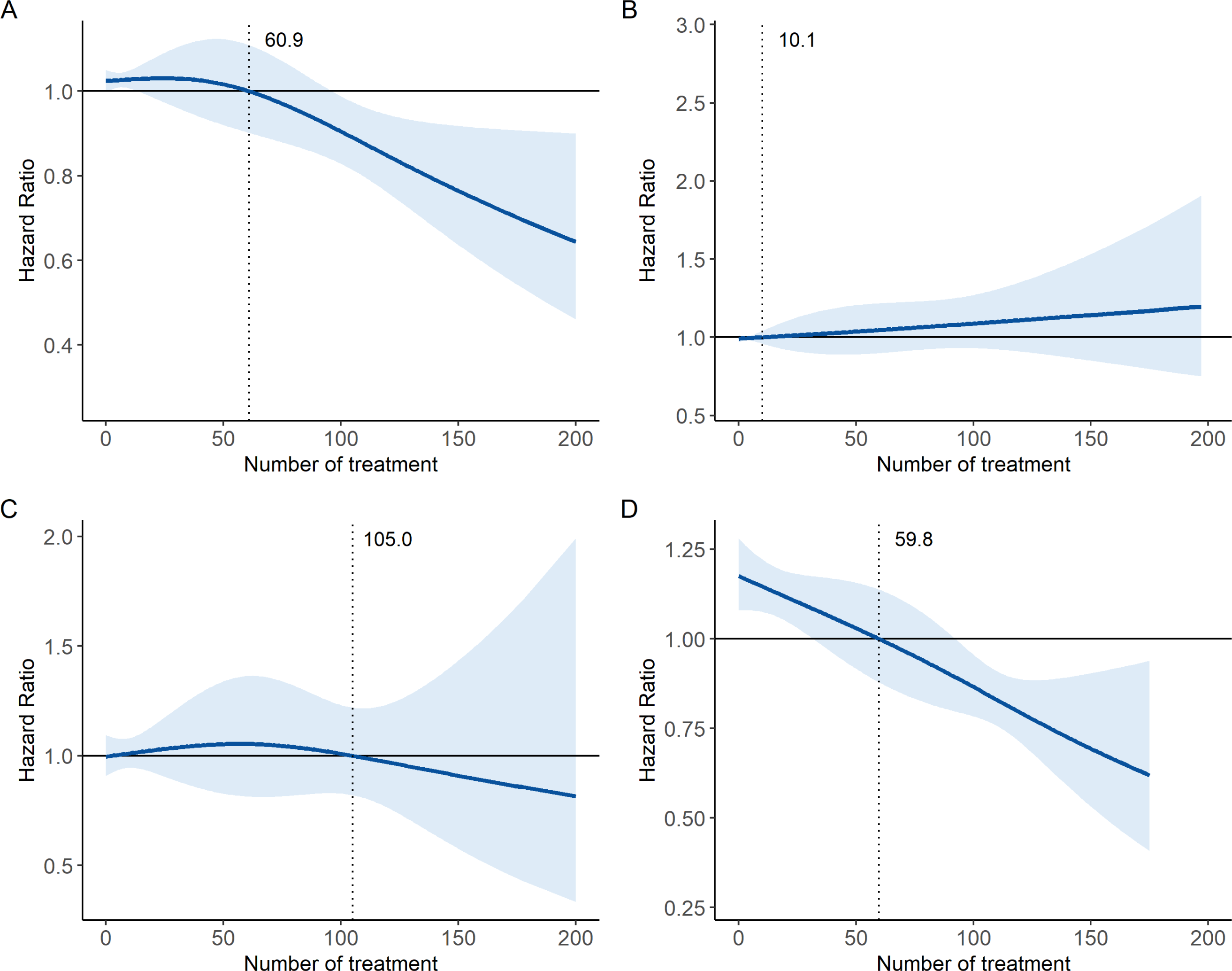
Restricted cubic spline plots of association between number of rehabilitation therap y and all-cause mortality in patients with stroke. (A) Total, (b) No disability, (c) mild to moderate disability, and (d) severe disability.

### Subgroup analysis by stoke type and sex

Table 3 summarizes the differences in the mortality risk according to the frequency of rehabilitation therapy for the two stroke types. In patients with ischemic stroke, there was no significant association between rehabilitation therapy and mortality, and this result was consistent regardless of disease severity. There was a significant dose-response association between rehabilitation therapy and mortality in patients with hemorrhagic stroke. When no rehabilitation group was used as a reference group, the HRs of lower and higher rehabilitation therapy groups were 0.81 (95% CI 0.67-0.98) and 0.60 (95% CI 0.49-0.74), respectively. In hemorrhagic stroke patients with disability, higher frequency of rehabilitation therapy was associated with reduced mortality rates compared to no rehabilitation therapy. (mild to moderate, HR= 0.46, 95% CI 0.24-0.87; severe, HR=0.51, 95% CI 0.38-0.68)

**Table 3.** Subgroup analysis of all-cause mortality by rehabilitation therapy frequency in patients with stroke according to stroke type.

| Stroke Type | Disability | Treatment | Stroke (n) | Mortality (n) | Person years | Mortality rate | Model 1 | p-value | Mode 2 | p-value | Mode 3 | p-value |
| --- | --- | --- | --- | --- | --- | --- | --- | --- | --- | --- | --- | --- |
| Ischemic | Total | No | 4573 | 1448 | 22363 | 64.75 | 1.00 (reference) |  | 1.00 (reference) |  | 1.00 (reference) |  |
| | | Yes ($\leq 40$) | 1443 | 419 | 6033 | 69.451 | 1.08 (0.97-1.21) | 0.115 | 1.09 (0.98-1.22) | 0.113 | 0.97 (0.86-1.10) | 0.574 |
| | | Yes ($> 40$) | 1206 | 389 | 5062 | 76.847 | 1.20 (1.07-1.34) | 0.002 | 1.31 (1.17-1.47) | <.001 | 1.04 (0.91-1.18) | 0.284 |
|  | None | No | 3406 | 988 | 16012 | 61.704 | 1.00 (reference) |  | 1.00 (reference) |  | 1.00 (reference) |  |
| | | Yes ($\leq 40$) | 947 | 250 | 3598 | 69.483 | 1.13 (0.98-1.30) | 0.085 | 1.13 (0.89-1.30) | 0.082 | 0.96 (0.82-1.13) | 0.639 |
| | | Yes ($> 40$) | 373 | 128 | 1221 | 104.832 | 1.71 (1.42-2.07) | <.001 | 1.37 (1.13-1.67) | 0.002 | 1.15 (0.93-1.40) | 0.194 |
|  | Mild to moderate | No | 666 | 199 | 3775 | 52.715 | 1.00 (reference) |  | 1.00 (reference) |  | 1.00 (reference) |  |
| | | Yes ($\leq 40$) | 288 | 69 | 1413 | 48.832 | 0.94 (0.71-1.23) | 0.642 | 1.03 (0.78-1.37) | 0.847 | 0.97 (0.72-1.30) | 0.824 |
| | | Yes ($> 40$) | 328 | 70 | 1410 | 49.645 | 0.96 (0.73-1.27) | 0.788 | 1.44 (1.08-1.92) | 0.013 | 1.31 (0.96-1.80) | 0.086 |
|  | Severe | No | 501 | 261 | 2576 | 78.568 | 1.00 (reference) |  | 1.00 (reference) |  | 1.00 (reference) |  |
| | | Yes ($\leq 40$) | 208 | 100 | 1022 | 95.238 | 0.97 (0.77-1.22) | 0.798 | 0.95 (0.75-1.19) | 0.647 | 1.00 (0.78-1.27) | 0.957 |
| | | Yes ($> 40$) | 505 | 191 | 2431 | 71.429 | 0.78 (0.65-0.94) | 0.010 | 0.85 (0.70-1.03) | 0.096 | 0.89 (0.72-1.10) | 0.288 |
| Hemorrhagic | Total | No | 1964 | 494 | 11069 | 44.629 | 1.00 (reference) |  | 1.00 (reference) |  | 1.00 (reference) |  |
| | | Yes ($\leq 40$) | 672 | 167 | 3506 | 47.633 | 1.06 (0.89-1.27) | 0.495 | 1.03 (0.86-1.23) | 0.781 | 0.81 (0.67-0.98) | 0.016 |
| | | Yes ($> 40$) | 898 | 184 | 4284 | 42.951 | 0.95 (0.80-1.12) | 0.547 | 1.12 (0.95-1.33) | 0.177 | 0.60 (0.49-0.74) | <.001 |
|  | None | No | 1525 | 308 | 8563 | 35.969 | 1.00 (reference) |  | 1.00 (reference) |  | 1.00 (reference) |  |
| | | Yes ($\leq 40$) | 437 | 89 | 2201 | 40.436 | 1.11 (0.88-1.40) | 0.395 | 0.89 (0.70-1.13) | 0.333 | 0.69 (0.54-0.89) | 0.004 |
| | | Yes ($> 40$) | 223 | 58 | 822 | 70.56 | 1.84 (1.34-2.45) | <.001 | 1.56 (1.18-2.06) | 0.002 | 0.93 (0.67-1.27) | 0.628 |
|  | Mild to moderate | No | 225 | 68 | 1350 | 3775 | 1.00 (reference) |  | 1.00 (reference) |  | 1.00 (reference) |  |
| | | Yes ($\leq 40$) | 113 | 23 | 681 | 1413 | 0.67 (0.42-1.09) | 0.105 | 0.94 (0.58-1.51) | 0.789 | 0.68 (0.39-1.19) | 0.178 |
| | | Yes ($> 40$) | 201 | 16 | 1001 | 1410 | 0.32 (0.18-0.55) | <.001 | 0.56 (0.32-0.99) | 0.047 | 0.46 (0.24-0.87) | 0.017 |
|  | Severe | No | 214 | 118 | 1156 | 102.076 | 1.00 (reference) |  | 1.00 (reference) |  | 1.00 (reference) |  |
| | | Yes ($\leq 40$) | 122 | 55 | 624 | 88.141 | 0.87 (0.62-1.23) | 0.436 | 1.00 (0.71-1.40) | 0.983 | 0.95 (0.68-1.34) | 0.783 |
| | | Yes ($> 40$) | 474 | 110 | 2462 | 44.679 | 0.44 (0.34-0.57) | <.001 | 0.53 (0.40-0.69) | <.001 | 0.51 (0.38-0.68) | <.001 |

Mortality rate is defined as the incidence of mortality per 1000 person-year
Model 1: unadjusted
Model 2: adjusted for age, sex, income level, insurance type, and area of residence
Model 3: adjusted for age, sex, income level, insurance type, area of residence, disability, comorbidities, lifestyle factors, and body mass index

In the subgroup analysis by sex, both male and female stroke patients with severe disabilities who received >40 rehabilitation therapy sessions showed lower mortality rates (Table S1).

### Factors associated with mortality after stroke according to disease severity

We also investigated the factors associated with mortality after stroke stratified by disease severity (Table 4). Overall, older age and male sex were associated with higher mortality rates, regardless of disease severity. The effect of age on mortality was less pronounced as disease severity increased. Majority of the sociodemographic factors, including income level, area of residence, type of insurance, type of stroke, and CCI, were not associated with mortality rates. Overall, pneumonia showed association with reduced mortality rate (HR=1.91, 95% CI 1.75-2.09), whereas dyslipidemia was associated with reduced mortality rate regardless of disease severity (HR=0.50, 95% CI 0.43-0.56).

**Table 4.**
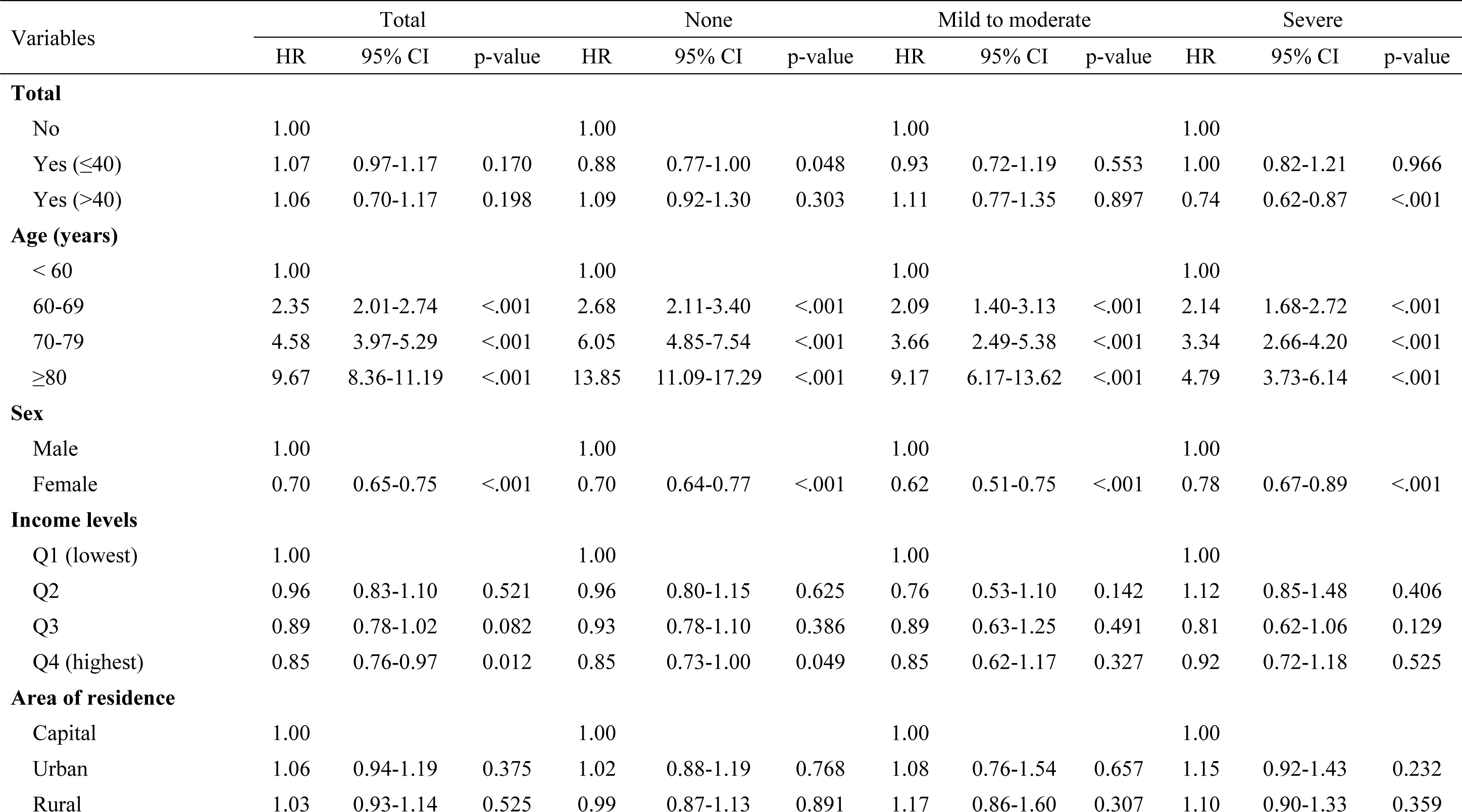
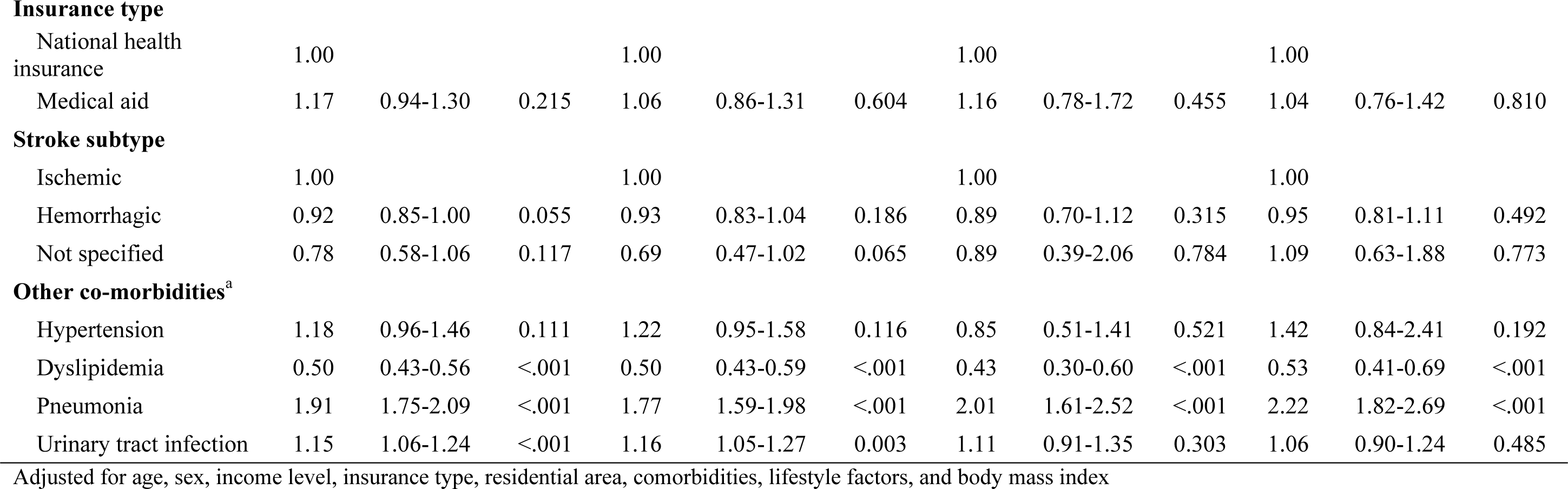
Adjusted Hazard Ratios for all-cause mortality after stroke according to disability severity.

## Discussion

In this nationwide population-based cohort study, we analyzed 10,974 patients with stroke to evaluate the association between rehabilitation therapy and long-term all-cause mortality. We observed a significantly reduced mortality rate in post-stroke patients who received a higher frequency of rehabilitation therapy than those who did not receive any therapy. For severely disabled post-stroke patients, the mortality rate was significantly reduced to 25%, and the inverse dose-response association between the number of rehabilitation therapy sessions and mortality rate was profound. In the subgroup analysis by stroke type, rehabilitation therapy was only associated with reduced mortality in patients with hemorrhagic stroke, demonstrating mortality risk reduction by approximately 50% in severely disabled patients. In patients with stroke, older age, male sex, and pneumonia were associated with an increased mortality risk, regardless of the degree of disability, whereas dyslipidemia was associated with reduced mortality risk. Sociodemographic factors including income level, area of residence, insurance type, and stroke type were not associated with mortality risk.

Numerous studies have supported the role of post-stroke rehabilitation in improving functional outcomes.^12, 24, 25^ However, few studies have reported conflicting results regarding the association between post-stroke rehabilitation therapy and mortality. Previous studies have shown that rehabilitation therapy is significantly associated with a lower post-stroke mortality risk. ^20, 21, 26^ Hou et al. reported that rehabilitation within the first three months after stroke was significantly associated with a lower risk of long-term mortality for 10 years after stroke.^16^ In addition, aggressive rehabilitation during the first year after ischemic stroke, with a high treatment frequency, has been shown to have a protective role in preventing stroke recurrence and mortality.^21^ On the other hand, non-significant association between rehabilitation therapy and mortality after stroke has also been reported. ^18, 19^. These discrepancies may originate from the diverse definitions of rehabilitation therapy, patient characteristics, and follow-up durations across the studies. In this study, among the various rehabilitation treatments, we focused on intensive physical therapy (claim code: MM105), the most important therapy for functional recovery after stroke in Korea, and evaluated the cumulative effects of rehabilitation therapy on long-term mortality in patients with stroke. Brain plasticity, which refers to modifications in the organization of neural components, has been reported to be responsible for adaptive changes in response to brain damage. ^27^ Previous studies have suggested that a critical period of enhanced plasticity occurs within the first 3-6 months after stroke. However, functional improvement was observed in the chronic phases. ^28-30^ Based on previous literature stating that significant recovery typically reaches its limit at six months, we evaluated the association between rehabilitation therapy within six months after stroke onset and all-cause mortality.

We found that increased frequency of rehabilitation therapy within six months was significantly associated with a reduced mortality rate after stroke. Furthermore, we also found that patients receiving more than 60 sessions of rehabilitation therapy showed a dose-response association between the number of rehabilitation therapies and mortality rate. According to the analysis of disability severity after stroke, the association between rehabilitation therapy and mortality rate was significant only in severely disabled patients. In addition, dose-response association between the number of rehabilitation therapies and mortality rate was more profound. Our results suggested the importance of volume of rehabilitation therapy on mortality rate and that rehabilitation therapy should be applied to patients with acute and subacute stroke, especially those with severe disability. In contrast, no significant association was observed between rehabilitation therapy and mortality in post-stroke patients with mild-to-moderate disabilities. Our findings are congruent with those of a previous cohort study, which suggested no correlation between the frequency of rehabilitation treatments and their outcomes in post-stroke patients with mild-to-moderate disabilities. ^31^ It has been suggested that among the various predictors of functional outcome in patients with stroke, stroke severity is considered one of the strongest prognostic factors.^32^ Mildly disabled stroke survivors are more likely to have less damage at the time of stroke. This means that for these patients, a lower proportion of recovery is needed to reach their prior neurological status, and functional recovery might be sufficient through perilesional reorganization near the stroke lesion. ^33^ This may be a possible reason for the non-significant association between rehabilitation therapy and mortality rate in these populations. On the other hand, among post-stroke patients with no disability, the mortality rate was significantly reduced in those who received ≤ 40 sessions of rehabilitation treatment. These patients may have a high level of interest and participation in healthcare management, thereby increasing their chances of survival.^34^

In the subgroup analysis according to stroke type, patients with hemorrhagic stroke who received a higher frequency of therapy had lower mortality rates than those without or with less rehabilitation therapy. However, there was no significant difference in the mortality rates according to rehabilitation therapy in patients with ischemic stroke. It has been suggested that hemorrhagic stroke is more severe than ischemic stroke and is associated with higher mortality rates, especially in the acute phase. ^35^ Despite the severe nature of hemorrhagic stroke, based on our results, a higher frequency of rehabilitation therapy was found to have beneficial effects on prognosis and mortality in patients who survived six months after hemorrhagic stroke. According to subgroup analysis by sex, significant reduction in mortality was observed in severely disabled stroke patients of both sexes, with about 10 % greater reduction in women than men. Post-stroke mortality in relation to rehabilitation therapy according to sex has rarely been investigated, and further studies are needed to corroborate our findings.

In this study, we also investigated the factors associated with mortality after stratifying stroke by disease severity. Older age, male sex, and pneumonia were associated with mortality in patients with stroke, which is consistent with the results of previous studies. ^36, 37^ On the other hand, dyslipidemia showed significant association with reduced mortality risk regardless of the severity of disability. Dyslipidemia is an important risk factor for ischemic stroke and statins are commonly prescribed to lower lipid levels. Some studies have suggested that statins have neuroprotective effects. ^38, 39^ In this study, we did not consider the use of statin in the analysis, and future studies considering statin use and lipid levels as predictors of post-stroke mortality are warranted. Interestingly, there was no significant association between socioeconomic status (SES), including income level, area of residence, and insurance type, and post-stroke mortality. Previous studies have shown that stroke incidence and mortality rates are higher in patients with low SES than in those with high SES.^40, 41^ In Korea, the NHIS offers healthcare coverage to almost every citizen. In addition to acute stroke care, post-stroke patients can receive intensive rehabilitation treatment for up to two years since the onset, either as inpatients or outpatients, at an economically affordable cost. This may explain the non-association between SES and poststroke mortality in the Korean population. The inverse association between number of rehabilitation therapies and mortality after stroke in our study may partially be attributed to the well-organized rehabilitation system covered by the NHIS in Korea, which raises awareness of the importance of both rehabilitation therapy and insurance coverage on the prognosis of patients with stroke.

This study had a few limitations that need to be considered. First, since this was a nationwide claims-based study, other treatments that the NHIS did not cover, such as speech therapy or cognitive therapy, were not reflected. Second, we focused on specific physical therapy (claim code: MM105)” among the various rehabilitation treatments. Although it is the most important therapy for functional recovery after stroke in Korea, it may have biased our results. Considering that various types of rehabilitation treatments and their cumulative effects on post-stroke mortality seemed inappropriate, we only focused on individualized 1:1 physical therapy in this study. Third, since stroke severity was not available from the NHIS data, disease severity was indirectly estimated using the NDR system. Fourth, detailed methods of rehabilitation therapy could differ across institutions, which could have affected the results.

## Data Availability

The data are available from the corresponding author on reasonable request.

## Acknowledgements

None

## References

1. Global, regional, and national burden of stroke and its risk factors, 1990-2019: a systematic analysis for the Global Burden of Disease Study 2019. Lancet Neurol 2021;20:795-820.

2. Korea. S. Causes of death statistics in 2021. 2022.

3. Jung SH. Stroke Rehabilitation Fact Sheet in Korea. Ann Rehabil Med 2022;46:1–8.

4. Rost NS, Brodtmann A, Pase MP, et al. Post-Stroke Cognitive Impairment and Dementia. Circ Res 2022;130:1252–1271.

5. Hackett ML, Köhler S, O’Brien JT, Mead GE. Neuropsychiatric outcomes of stroke. Lancet Neurol 2014;13:525–534.

6. Hatem SM, Saussez G, Della Faille M, et al. Rehabilitation of Motor Function after Stroke: A Multiple Systematic Review Focused on Techniques to Stimulate Upper Extremity Recovery. Front Hum Neurosci 2016;10:442.

7. Cha YJ. The Economic Burden of Stroke Based on South Korea’s National Health Insurance Claims Database. Int J Health Policy Manag 2018;7:904–909.

8. Avan A, Digaleh H, Di Napoli M, et al. Socioeconomic status and stroke incidence, prevalence, mortality, and worldwide burden: an ecological analysis from the Global Burden of Disease Study 2017. BMC Medicine 2019;17:191.

9. Kim Y-H, Han TR, Jung HY, et al. Clinical Practice Guideline for Stroke Rehabilitation in Korea. Brain Neurorehabil 2009;2:1–38.

10. Kuptniratsaikul V, Kovindha A, Piravej K, Dajpratham P. First-Year Outcomes after Stroke Rehabilitation: A Multicenter Study in Thailand. ISRN Rehabilitation 2013;2013:595318.

11. Park D, Son KJ, Kim HS. Chronic Phase Survival Rate in Stroke Patients With Severe Functional Limitations According to the Frequency of Rehabilitation Treatment. Arch Phys Med Rehabil 2023;104:251–259.

12. Yagi M, Yasunaga H, Matsui H, et al. Impact of Rehabilitation on Outcomes in Patients With Ischemic Stroke: A Nationwide Retrospective Cohort Study in Japan. Stroke 2017;48:740–746.

13. Aprile I, Di Stasio E, Romitelli F, et al. Effects of rehabilitation on quality of life in patients with chronic stroke. Brain Injury 2008;22:451–456.

14. Alawieh A, Zhao J, Feng W. Factors affecting post-stroke motor recovery: Implications on neurotherapy after brain injury. Behav Brain Res 2018;340:94–101.

15. Jørgensen HS, Nakayama H, Raaschou HO, Larsen K, Hübbe P, Olsen TS. The effect of a stroke unit: reductions in mortality, discharge rate to nursing home, length of hospital stay, and cost. A community-based study. Stroke 1995;26:1178–1182.

16. Hou W-H, Ni C-H, Li C-Y, Tsai P-S, Lin L-F, Shen H-N. Stroke Rehabilitation and Risk of Mortality: A Population-Based Cohort Study Stratified by Age and Gender. Journal of Stroke and Cerebrovascular Diseases 2015;24:1414–1422.

17. Jørgensen HS, Kammersgaard LP, Nakayama H, et al. Treatment and rehabilitation on a stroke unit improves 5-year survival. A community-based study. Stroke 1999;30:930–933.

18. Park D, Son KJ, Kim JH, Kim HS. Effect of the Frequency of Rehabilitation Treatments on the Long-Term Mortality of Stroke Survivors with Mild-to-Moderate Disabilities under the Korean National Health Insurance Service System. Healthcare 2023;11:1587.

19. Efficacy and safety of very early mobilisation within 24 h of stroke onset (AVERT): a randomised controlled trial. Lancet 2015;386:46-55.

20. Hsieh CY, Huang HC, Wu DP, Li CY, Chiu MJ, Sung SF. Effect of Rehabilitation Intensity on Mortality Risk After Stroke. Arch Phys Med Rehabil 2018;99:1042–1048.e1046.

21. Cheng YY, Shu JH, Hsu HC, et al. The Impact of Rehabilitation Frequencies in the First Year after Stroke on the Risk of Recurrent Stroke and Mortality. J Stroke Cerebrovasc Dis 2017;26:2755–2762.

22. Kim YW. Update on Stroke Rehabilitation in Motor Impairment. Brain Neurorehabil 2022;15.

23. Branco JP, Oliveira S, Sargento-Freitas J, Laíns J, Pinheiro J. Assessing functional recovery in the first six months after acute ischemic stroke: a prospective, observational study. Eur J Phys Rehabil Med 2019;55:1–7.

24. Van Peppen RP, Kwakkel G, Wood-Dauphinee S, Hendriks HJ, Van der Wees PJ, Dekker J. The impact of physical therapy on functional outcomes after stroke: what’s the evidence? Clin Rehabil 2004;18:833–862.

25. McGlinchey MP, James J, McKevitt C, Douiri A, Sackley C. The effect of rehabilitation interventions on physical function and immobility-related complications in severe stroke: a systematic review. BMJ Open 2020;10:e033642.

26. Hu GC, Hsu CY, Yu HK, Chen JP, Chang YJ, Chien KL. Association between the volume of inpatient rehabilitation therapy and the risk of all-cause and cardiovascular mortality in patients with ischemic stroke. Arch Phys Med Rehabil 2014;95:269–275.

27. Alia C, Spalletti C, Lai S, et al. Neuroplastic Changes Following Brain Ischemia and their Contribution to Stroke Recovery: Novel Approaches in Neurorehabilitation. Front Cell Neurosci 2017;11:76.

28. Ballester BR, Maier M, Duff A, et al. A critical time window for recovery extends beyond one-year post-stroke. J Neurophysiol 2019;122:350–357.

29. Dromerick AW, Geed S, Barth J, et al. Critical Period After Stroke Study (CPASS): A phase II clinical trial testing an optimal time for motor recovery after stroke in humans. Proc Natl Acad Sci U S A 2021;118.

30. Grefkes C, Fink GR. Recovery from stroke: current concepts and future perspectives. Neurological Research and Practice 2020;2:17.

31. Park D, Son KJ, Kim JH, Kim HS. Effect of the Frequency of Rehabilitation Treatments on the Long-Term Mortality of Stroke Survivors with Mild-to-Moderate Disabilities under the Korean National Health Insurance Service System. Healthcare (Basel) 2023;11.

32. Thijs VN, Lansberg MG, Beaulieu C, Marks MP, Moseley ME, Albers GW. Is early ischemic lesion volume on diffusion-weighted imaging an independent predictor of stroke outcome? A multivariable analysis. Stroke 2000;31:2597–2602.

33. Lee HH, Kim DY, Sohn MK, et al. Revisiting the Proportional Recovery Model in View of the Ceiling Effect of Fugl-Meyer Assessment. Stroke 2021;52:3167–3175.

34. Yang CP, Cheng HM, Lu MC, Lang HC. Association between continuity of care and long-term mortality in Taiwanese first-ever stroke survivors: An 8-year cohort study. PLoS One 2019;14:e0216495.

35. Andersen KK, Olsen TS, Dehlendorff C, Kammersgaard LP. Hemorrhagic and ischemic strokes compared: stroke severity, mortality, and risk factors. Stroke 2009;40:2068–2072.

36. Yoo DY, Choi JK, Baek CY, Shin JB. Impact of intensive rehabilitation on long-term prognosis after stroke: A Korean nationwide retrospective cohort study. Medicine (Baltimore) 2022;101:e30827.

37. Wilson RD. Mortality and cost of pneumonia after stroke for different risk groups. J Stroke Cerebrovasc Dis 2012;21:61–67.

38. Vaughan CJ, Delanty N. Neuroprotective properties of statins in cerebral ischemia and stroke. Stroke 1999;30:1969–1973.

39. Fisher M, Moonis M. Neuroprotective effects of statins: evidence from preclinical and clinical studies. Curr Treat Options Cardiovasc Med 2012;14:252–259.

40. Brown AF, Liang LJ, Vassar SD, et al. Neighborhood socioeconomic disadvantage and mortality after stroke. Neurology 2013;80:520–527.

41. Addo J, Ayerbe L, Mohan KM, et al. Socioeconomic status and stroke: an updated review. Stroke 2012;43:1186–1191.

